# Digital twin reconstruction of ventricular repolarisation identifies regional causes of T-wave abnormalities in hypertrophic cardiomyopathy

**DOI:** 10.64898/2026.06.15.26355737

**Authors:** James A Coleman, Julia Camps, Abdallah I Hasaballa, Rina Ariga, Betty Raman, Iacopo Olivotto, Hugh Watkins, Alfonso Bueno-Orovio

## Abstract

**Background:** Abnormal ventricular repolarisation in hypertrophic cardiomyopathy (HCM) may predispose patients to lethal arrhythmias, but repolarisation in HCM remains poorly spatially characterised. Integrating spatial cardiac magnetic resonance with ECG data has the potential to map electrical function throughout the ventricles. This study applied novel digital twin inverse ECG methods to map repolarisation patterns underlying abnormal T-waves in HCM.

**Methods:** Patient-specific full ventricular electrophysiological models were iteratively refined to match the patient 12-lead ECG. Data from 32 healthy volunteers and 69 HCM patients were analysed, with inferred substrates incorporating activation times, repolarisation times, and rate-corrected action potential durations (APD_c_s). Patients were stratified by T-wave phenotype to identify distinct spatial repolarisation signatures associated with different ECG presentations.

**Results:** Clinical 12-lead ECGs were accurately reproduced by the inferred ventricular models in 95 of 101 cases. Healthy volunteers (N=30) and HCM patients with normal T-waves (N=33) were characterised by apex-to-base APD_c_ gradients of 60 ms (40–80) and 60 ms (30–80), respectively. HCM patients with V1-V3 T-wave abnormalities (N=6) had attenuated apex-to-base APD_c_ gradients of 30 ms (-20–40) driven by apical-to-mid anterior APD_c_ prolongation, greatest at the apical segment (ΔAPD_c_ vs. healthy: 54 ms; 95% CI: 23–84 ms). HCM patients with V4-V6 T wave abnormalities (N=21) had reversed apex-to-base APD_c_ gradients of -20 ms (-40–0) driven by apical-to-mid APD_c_ prolongation, most severe at the apical segment (ΔAPD_c_ vs. healthy: 94 ms; 95% CI: 74–120 ms). Despite significant APD_c_ prolongation, only 4 of 69 HCM patients had QT_c_ > 480 ms, due to masking by the intrinsically healthy longer APD_c_s at the ventricular base.

**Conclusions:** Distinct spatial distributions of APD_c_ prolongation, not necessarily mirroring the distribution of hypertrophy, underlie different ECG repolarisation phenotypes in HCM and may be missed by the QT_c_ interval.

**Clinical Perspective:** *What is Known:*

- T-wave abnormalities on ECG occur in 50% of HCM patients, but the drivers of these ECG changes remain only partially understood.
- Abnormal prolonged repolarisation is thought to occur in HCM, but QT_c_ > 480 ms on ECG occurs only in 13% of HCM patients.

*What the Study Adds:*

- This study uses a simulation-based inference framework to reconstruct patient-specific spatially resolved electrophysiological substrates in HCM from routine 12-lead ECG and CMR.
- By comparing to healthy volunteers, the framework identifies prolongation of ventricular repolarisation in mid-to-apical regions underlying T-wave abnormalities in HCM, indicating a regionally impaired electrical substrate.
- The framework identifies that the QT_c_ interval fails to capture regional prolongation of repolarisation in HCM, particularly in mid-to-apical ventricular regions.

## 1. Introduction

Risk stratification in hypertrophic cardiomyopathy (HCM) remains a significant clinical challenge (Maron et al., 2019). Lethal arrhythmias in HCM are thought to arise from an electrophysiological substrate including both impaired ventricular activation and repolarisation (Johnson et al., 2011; Joy et al., 2024). Whilst progress has been made in the spatial characterisation of abnormal activation, primarily through late gadolinium enhancement cardiac magnetic resonance (CMR) imaging of fibrosis (Weng et al., 2016), the spatial understanding of repolarisation abnormalities in HCM remains incomplete. Consequently, repolarisation is not currently considered in clinical risk-stratification guidelines (Arbelo et al., 2023; Ommen et al., 2020).

Notably, abnormal repolarisation constitutes the most common ECG abnormality in HCM, with T-wave inversion present in the majority of pathological ECGs (Dores et al., 2023). Despite its high prevalence, the distribution of T-wave inversion among leads is highly clinically heterogeneous, even within the same structural HCM phenotype (Park et al., 2018); the extent of lead involvement can vary widely, from a single lead to all twelve. However, the precise myocardial substrate underlying these distinct ECG patterns is as yet unknown.

At the cellular scale, HCM may be characterised by prolonged action potential duration (APD), as shown in patch-clamp studies of human HCM myectomy samples (Coppini et al., 2013). Organ-scale computer models of these repolarisation changes showed that they can give rise to T-wave inversions and QT interval prolongation (Coleman et al., 2024a; Coppini et al., 2019; Lyon et al., 2018b). One apparent contradiction, however, is that despite T-wave inversions affecting 50% of HCM patients, clinically relevant QT_c_ prolongation (>480 ms) is only reported in 13% of cases (Johnson et al., 2011; Joy et al., 2024; Zeppenfeld et al., 2022). This discrepancy is particularly evident in apical HCM, where over 90% of cases exhibit T-wave inversion, yet on average QT_c_ intervals remain within the normal range (Eriksson et al., 2002). Therefore, it is unclear how T-wave inversion may occur without QT_c_ prolongation in so many, if APD prolongation underlies T-wave inversions in HCM.

Personalised simulation-based inference is emerging as a promising method to non-invasively assess ventricular electrophysiology, including repolarisation (Campos et al., 2024; Camps et al., 2025; Sánchez et al., 2025). Briefly, these methodologies integrate CMR and ECG data with computer simulations, to infer myocardial conduction patterns that are consistent with the patient ventricular anatomy and body-surface potential recordings. This is achieved by the iterative refinement of parameters within three-dimensional electrophysiology simulations, to reconstruct the patient-specific electrical substrate underlying their observed ECG. We have recently developed and evaluated a simulation-based inference pipeline that can reconstruct with state-of-the-art accuracy ventricular activation and repolarisation patterns in both healthy and diseased cases (Coleman et al., 2025).

The present study therefore aimed to adapt this simulation-based inference approach for application to a cohort of 69 HCM patients and 32 healthy volunteers, to investigate patient-specific causes of T-wave abnormalities and effects on the QT interval. We hypothesised that regional APD prolongation may underlie abnormal repolarisation patterns and T-wave morphology in HCM. This approach is expected to offer non-invasive and patient-specific insights into repolarisation in HCM, which may hold potential for future application to arrhythmic risk assessment and pharmacologic therapy optimisation (Chauhan et al., 2006; Coleman et al., 2024a; Corral-Acero et al., 2020; Yamazaki et al., 2022).

## 2. Methods

### 2.1 Study population

This study included 69 HCM patients and 32 healthy volunteers. Participants were drawn from a previously described prospective cohort of 85 HCM and 38 healthy volunteers recruited from the University of Oxford Inherited Cardiac Conditions clinic, John Radcliffe Hospital, Oxford, UK (REC ref 12/LO/1979) (Lyon et al., 2018a). HCM diagnosis was made based on the presence of a pathogenic mutation in a known sarcomeric gene or, in the absence of an identified mutation, HCM was defined as left ventricular (LV) hypertrophy (≥15 mm) not originating from another cause. Gene positive patients without hypertrophy (G+LVH-) were included in the study. Age- and gender-matched healthy volunteers were non-smokers without cardiovascular disease, hypertension, diabetes, or family history of cardiomyopathy or sudden cardiac death. Individuals were excluded based on availability of sufficiently high-quality CMR to enable accurate ventricular anatomical reconstruction.

CMR images were acquired using long- and short-axis cine MRI at 3T (TIM Trio, Siemens) (Lyon et al., 2018b), and analysed using cmr42© (Circle Cardiovascular Imaging, Calgary, Canada). A standard 10 s strip of 12-lead resting digital ECG was recorded for each participant (Burdick 8500, Glasgow, UK). All patients were in sinus rhythm during ECG acquisition.

### 2.2 Subgroup analysis by ECG T-wave phenotype

For the analysis of patient electrophysiological substrates, the 69 HCM patients introduced in Section 2.1 were classified into repolarisation subgroups based on the presence and distribution of T-wave abnormalities across the 12-lead ECG (see later Fig. 3). These HCM subgroups were defined as:

- *HCM normal T-waves (N=33):* HCM patients without abnormal T-waves.
- *HCM V1-V3 T-waves affected (N=8):* HCM patients with flat or inverted T-waves in ≥2 contiguous leads from V1-V3 excluding isolated V1 variants, and without T-wave abnormalities in V5-V6.
- *HCM V4-V6 T-waves affected (N=23):* HCM patients with flat or inverted T-waves in ≥2 contiguous leads from V4-V6, often also affecting leads I, II, aVL, aVR and V2-V3.
- *HCM with other T-wave abnormalities (N=5):* HCM patients with any other T-wave abnormalities, such as a globally abnormal T-wave shape, or T-wave abnormalities confined to limb leads.

The different HCM subgroups were compared with *healthy volunteers (N=32)*.

### 2.3 Data preprocessing

Patient-specific biventricular anatomical models were reconstructed from CMR images in previous work (Lyon et al., 2018b; Zacur et al., 2017). Briefly, after slice alignment and manual contour delineation, heart surfaces were generated through iterative remeshing, smoothing and fitting to the contours. Patient-specific volumetric biventricular meshes were then created using a flood filling method.

Beats from the 10 s ECG were detected and aligned using R peaks. Beats were assessed for consistency and any beats with artefacts were excluded. The median waveform was then computed for each lead from the remaining beats. This produced a processed clinical 12-lead ECG consisting of a single beat archetype. QRS complex and T-wave subsets of the processed ECG were manually selected for use in subsequent analysis. The QRS segment was taken from the onset of the Q-wave to the end of the S-wave in the lead with the longest and most clearly defined QRS complex. The T-wave segment was taken from the end of the S-wave to the end of the T-wave in the lead with the longest and most discernible T-wave.

### 2.4 Electrophysiological substrate inference

#### 2.4.1 Overview

Ventricular electrophysiology simulations were used to infer electrical substrates consistent with each patient biventricular anatomy and corresponding 12-lead ECG (Fig. 1A). The method simulates many virtual heartbeats with varying electrical properties and generates their associated 12-lead ECGs, iteratively refining the electrical parameters until close agreement between the simulated and clinically recorded ECGs. The inference algorithm and parameterisation of substrates, as developed and evaluated in previous work (Coleman et al., 2025), are described in detail in Supplementary Section 1.

**Figure 1.**
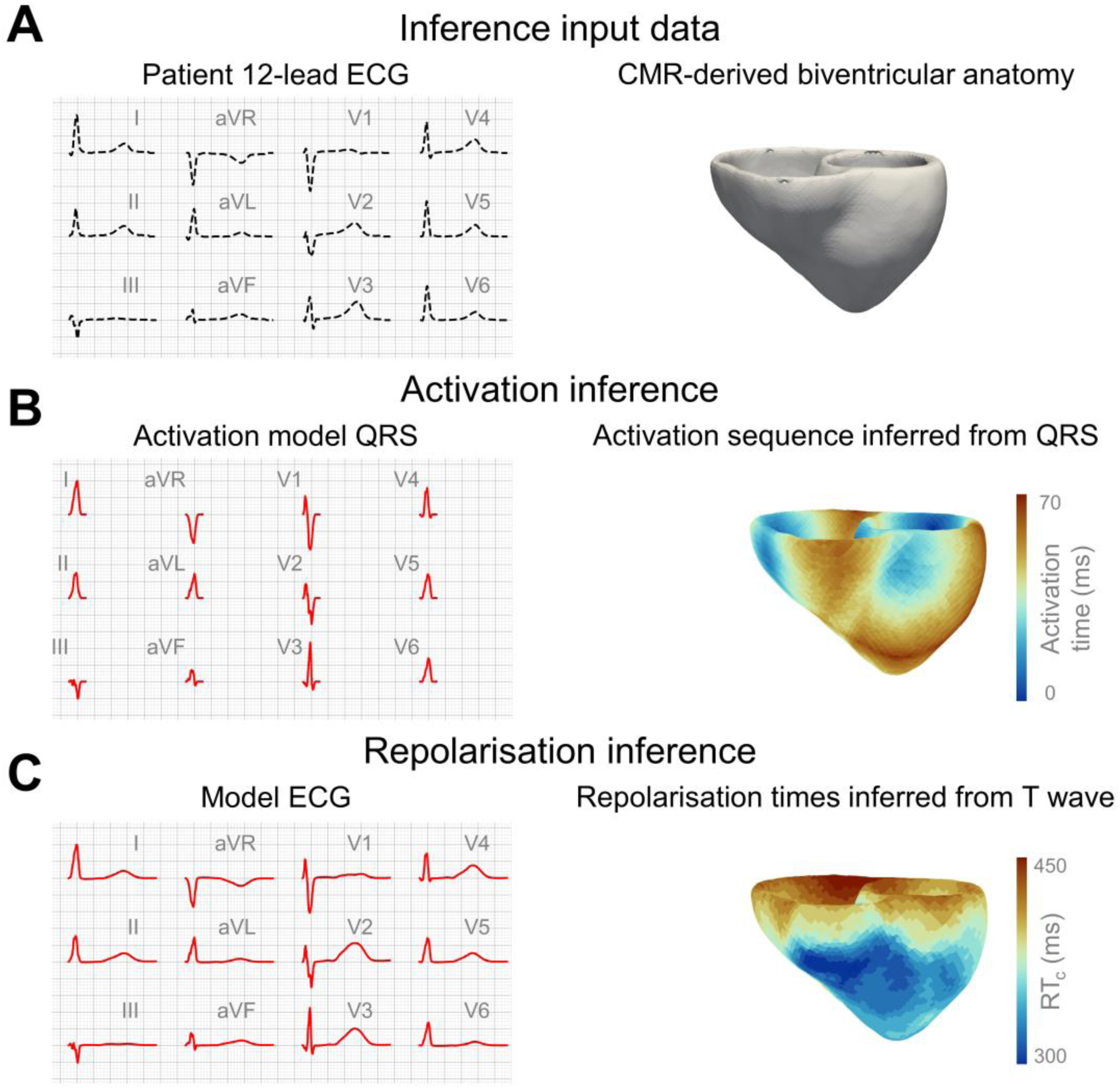
Inference methodology overview. (A) Input data to the inference process consists of the patient’s 12-lead ECG (used as target ECG) and CMR-derived biventricular anatomy. (B) Activation inference involves fitting ventricular activation properties by matching simulated QRS complexes to their target counterparts. (C) Repolarisation inference involves fitting ventricular repolarisation properties by matching simulated T-waves to their targets.

Because QRS and T-wave formation arise from distinct physiological processes (ventricular activation and repolarisation, respectively), the method first infers activation properties consistent with the patient’s QRS complexes (Fig. 1B), then infers repolarisation properties consistent with T-waves (Fig. 1C). This process yields a patient-specific electrophysiological substrate underlying their corresponding ECG, including inferred ventricular activation times, repolarisation times, and APDs at 90% repolarisation.

Regularisation, as commonly employed in ECG imaging, was used to impose smooth and physiologically plausible repolarisation patterns. The regularisation strength was tuned as described in Supplementary Section 1.3.2, with the full cohort analysis repeated at stronger regularisation levels to confirm stability and robustness (see Supplementary Section 2.5).

To enable comparisons between patients with differing heart rates, Bazett rate correction was applied to inferred repolarisation properties, yielding rate-corrected repolarisation times (RT_c_) and rate-corrected APD_90_ values (APD_c_) at 90% repolarisation.

#### 2.4.2 Validation

The methodology has previously been developed and validated against a range of control and HCM benchmark datasets, showing that ground-truth ventricular properties can be accurately reconstructed from the 12-lead ECG and biventricular anatomy (Camps et al., 2021; Coleman et al., 2025). For the present study, validation was repeated using the same 18 benchmark cases reported in our earlier work (Coleman et al., 2025). Inferred activation times showed strong agreement with ground-truth values (Spearman’s *r* = 0.72, range 0.50-0.80), with similar performance observed for inferred RT_c_s (Spearman’s *r* = 0.67, 0.56-0.77) (Supplementary Fig. S1). Validation against established repolarisation gradients was also performed (see Section 3.3).

#### 2.4.3 Modelling and simulation

Personalised simulations of electrophysiological substrates used the Eikonal formulation to model ventricular activation (Wallman et al., 2012), and the fast repolarisation surrogate model previously described in (Coleman et al., 2025) to model ventricular repolarisation. Briefly, the repolarisation surrogate model approximates reaction-diffusion behaviour by first assigning action potentials throughout the ventricular mesh using a precomputed lookup table derived from the human ToR-ORd action potential model (Tomek et al., 2019), then spatial smoothing to approximate electrical diffusion. Further details are provided in Supplementary Section 1.4.

Membrane potentials generated by the surrogate model were used to compute simulated ECGs, derived from 10 electrodes placed at rule-based locations around the ventricles, computed using a pseudo-ECG integral method (Bishop and Plank, 2011; Camps et al., 2025).

### 2.5 Statistical analysis

Data are expressed as mean ± standard deviation or median (interquartile range, IQR), as appropriate. Continuous clinical characteristics were compared using independent t-tests with Welch’s correction. Categorical clinical characteristics are expressed as percentages and were compared with chi-squared tests. Regional repolarisation differences between subgroups were analysed using bootstrapping among the American Heart Association 17 segments. Statistical significance was defined as *p* < 0.05, or when bootstrapping 95% confidence intervals excluded zero. Spearman’s rank correlation coefficients were used to assess similarity between activation and repolarisation sequences and ECG morphologies. Statistical analysis was performed using the scipy.stats Python library.

## 3. Results

### 3.1 Clinical characteristics

This study included 69 HCM patients and 32 healthy volunteers (Table 1).

**Table 1.** Clinical characteristics of healthy volunteers vs. HCM patients. *p<0.05

|  | Healthy (N=32) | HCM (N=69) | p value |
| --- | --- | --- | --- |
| **Demographics** |  |  |  |
| Age (years) | 46 ± 15 | 45 ± 15 | 0.708 |
| Male (%) | 70 | 67 | 0.927 |
| Body mass index (kg/m<sup>2</sup>) | 25 ± 4 | 26 ± 4 | 0.192 |
| Body surface area (m<sup>2</sup>) | 1.9 ± 0.2 | 1.9 ± 0.2 | 0.869 |
| **CMR** |  |  |  |
| LV End-diastolic volume (mL) | 160 ± 36 | 150 ± 32 | 0.358 |
| LV End-systolic volume (mL) | 52 ± 18 | 41 ± 16 | 0.007* |
| LV Ejection fraction (%) | 68 ± 6 | 73 ± 8 | <0.001* |
| LV Mass index (g/m<sup>2</sup>) | 55 ± 11 | 70 ± 25 | <0.001* |
| Max LV thickness (mm) | 11 ± 1 | 19 ± 6 | <0.001* |
| **ECG** |  |  |  |
| Heart rate (bpm) | 59 ± 11 | 58 ± 11 | 0.69 |
| QRS axis (°) | 32 ± 38 | 16 ± 47 | 0.082 |
| QRS duration (ms) | 94 ± 14 | 98 ± 13 | 0.185 |
| T wave axis (°) | 34 ± 17 | 42 ± 72 | 0.388 |
| QT<sub>c</sub> interval | 410 ± 20 | 430 ± 30 | <0.001* |

Age, sex, body mass index, and body surface area were similar between groups. Compared with healthy volunteers, HCM patients had lower LV end-systolic volume (41±16 vs. 52±18 mL; p=0.007) and higher LV ejection fraction (73±8% vs. 68±6%; p<0.001). In addition, HCM patients had a higher LV mass index (70±25 vs. 55±11 g/m^2^; p<0.001), greater maximal LV wall thickness (19±6 vs. 11±1 mm; p<0.001), as well as moderately prolonged QT_c_ interval (410±20 vs. 430±30 ms; p<0.001). Resting LV outflow tract obstruction (>30 mmHg) was present in 12% of HCM patients.

### 3.2 Patient ECGs are accurately reproduced by digital twins

In total, 101 personalised biventricular electrophysiological substrates were reconstructed from clinical ECG and CMR data. These comprised ventricular activation times, RT_c_s, APD_c_s, and the corresponding simulated ECGs. The patient-specific biventricular anatomies are shown in Fig. 2A.

**Figure 2.**
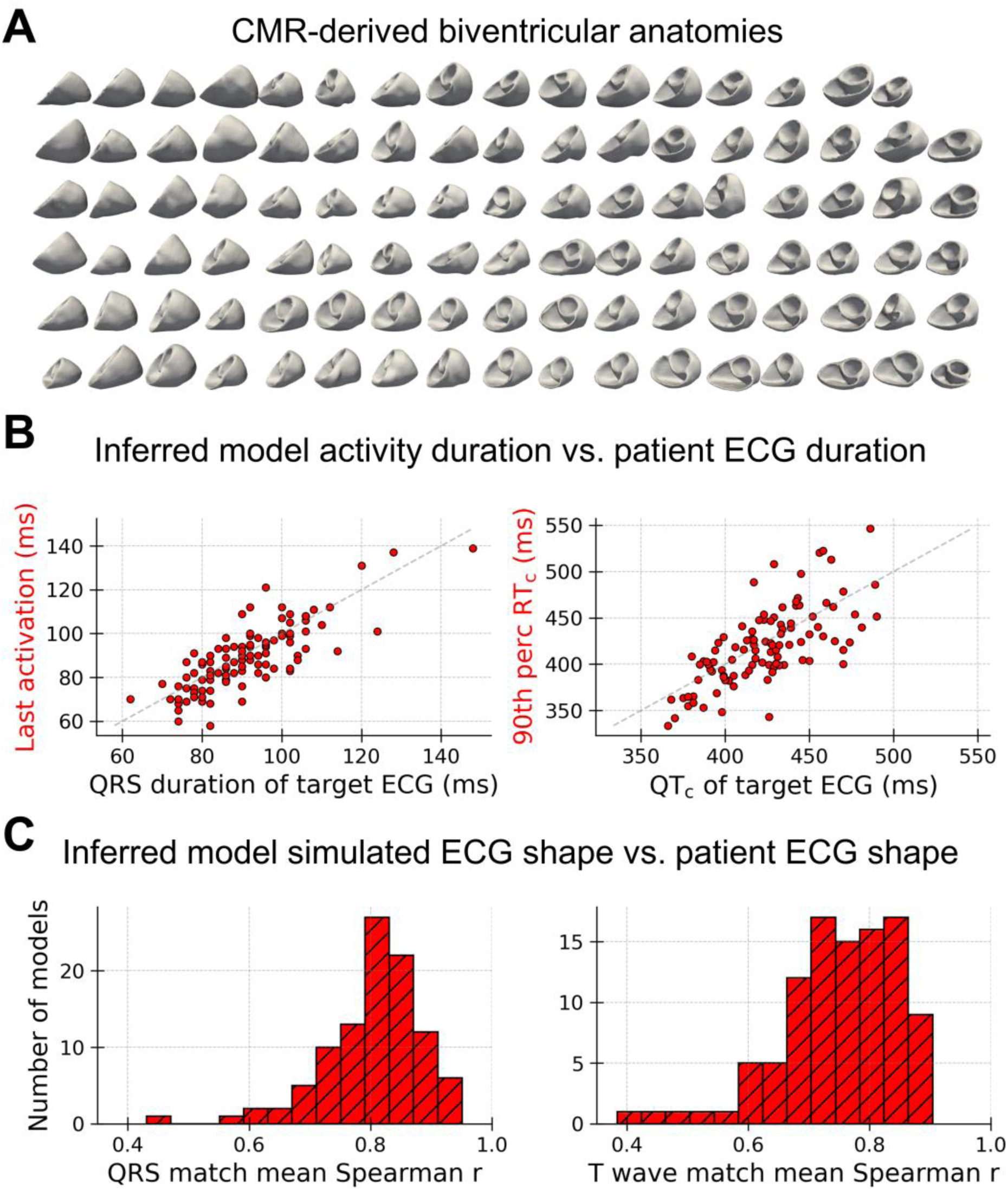
Overview of the 101 inferred biventricular electrophysiological substrates. (A) Patient-specific three-dimensional anatomical ventricular models derived from CMR imaging. (B) Latest activation time of the inferred ventricular model vs. QRS duration of the patient ECG (left), and 90^th^ percentile RT_c_ of the inferred ventricular model vs. QT_c_ of the patient ECG (right). (C) Quantitative assessment of shape similarity between simulated and clinical signals for QRS complexes (left) and T-waves (right), evaluated using mean Spearman’s r coefficient across the 12 leads.

The inferred timescales of ventricular activation and repolarisation were broadly consistent with the QRS duration and QT_c_ interval of the respective clinical ECGs (Fig. 2B), indicating that the reconstructed substrates captured the timing of ventricular depolarisation and repolarisation observed clinically. Simulated QRS complexes closely matched in morphology their clinical counterparts (12-lead average Spearman *r* = 0.82, IQR 0.76-0.86) (Fig. 2C, left). Similarly, simulated T-waves showed good morphological agreement with clinical T-waves (12-lead average Spearman’s *r* = 0.76, IQR 0.70-0.83) (Fig. 2C, right).

Overall, the accuracy of the reconstructed 12-lead ECGs was consistent with previous quantitative results obtained using synthetic benchmarks of human ventricular function across both control and HCM populations (Coleman et al., 2025). These findings further demonstrate the ability of the inferred models to reproduce QRS and T-wave morphologies across a heterogeneous clinical cohort, with good agreement. Only 6 of the 101 reconstructed cases (2 healthy volunteers, 2 cases with V1-V3 affected, and 2 cases with V4-V6 affected) showed weaker correspondence with their respective clinical ECG phenotype (12-lead average Spearman’s *r* < 0.6), and were therefore excluded from subsequent subgroup analysis (see Section 3.4).

### 3.3 Validation using established repolarisation gradients

Consistent with established human LV electrophysiology data (Glukhov et al., 2010; Srinivasan et al., 2019; Taggart et al., 2001), inferred APD_c_s were longer in LV endocardium than epicardium in both healthy volunteers (endo- vs. epicardial ΔAPD_c_: 9 ms; 95% CI: 7-11 ms) and HCM patients (endo- vs. epicardial ΔAPD_c_: 8 ms; 95% CI: 6-9 ms). These modest differences align with in vivo human observations using plunge electrodes, where any transmural repolarisation gradients were nominally small (<10 ms) (Taggart et al., 2001). When transmural gradients were analysed segmentally across the entire cohort, statistically significant effects were identified in basal lateral segments (Supplementary Fig. S2). This was greatest at the basal anterolateral segment (endo- vs. epicardial ΔAPD_c_: 14 ms; 95% CI: 2-26 ms), possibly influenced by basal wall thickness and electrode proximity. In addition to transmural gradients, inferred APD_c_s were evaluated against established apex-to-base repolarisation patterns. Consistent with prior human ECG imaging studies (Ramanathan et al., 2006), healthy volunteers exhibited apex-to-base APD_c_ gradients of 60 ms (40-80).

### 3.4 HCM T-wave abnormalities are driven by regional prolongation of repolarisation

When inferred ventricular electrophysiological substrates were analysed by patient subgroup (see Section 2.2), distinct APD_c_ patterns were observed corresponding to different T-wave presentations. Representative cases are provided in Fig. 3, with segmental summary data presented in Fig. 4A. Uncertainty in these representative cases shown in Fig. 3 was considered by analysing inter-run variability and regularisation effects, as shown in Supplementary Figs S4 & S5, respectively.

**Figure 3.**
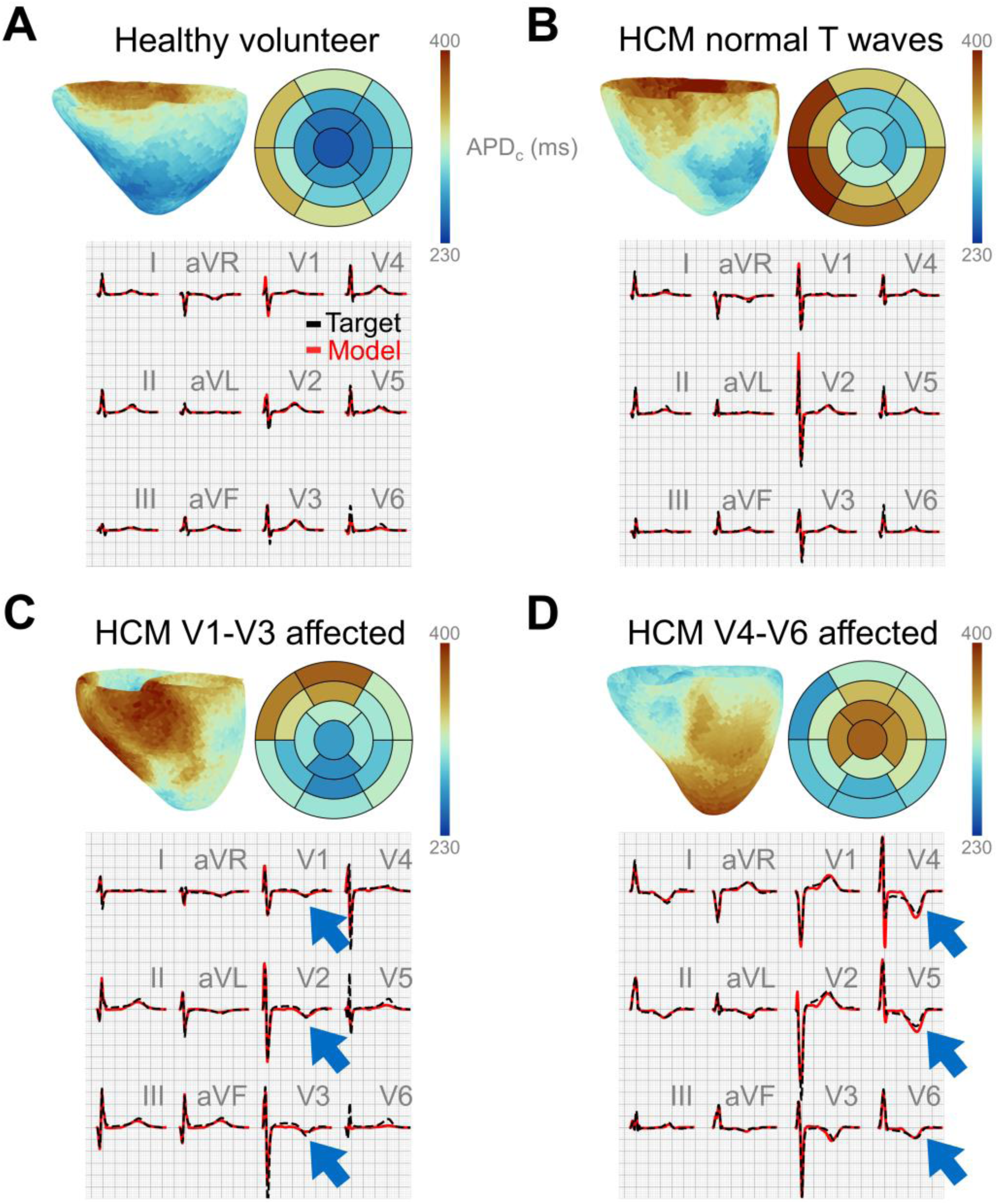
Representative inferred ventricular repolarisation phenotypes. Inferred ventricular APD_c_s shown from an anterior view, together with the corresponding APD_c_ bullseye plot and comparisons between inferred model and clinical 12-lead ECGs. Representative cases are shown for: (A) healthy volunteers, (B) HCM patients with normal T-waves, (C) HCM patients with abnormal T-waves in V1-V3, and (D) HCM patients with abnormal T-waves in V4-V6.

**Figure 4.**
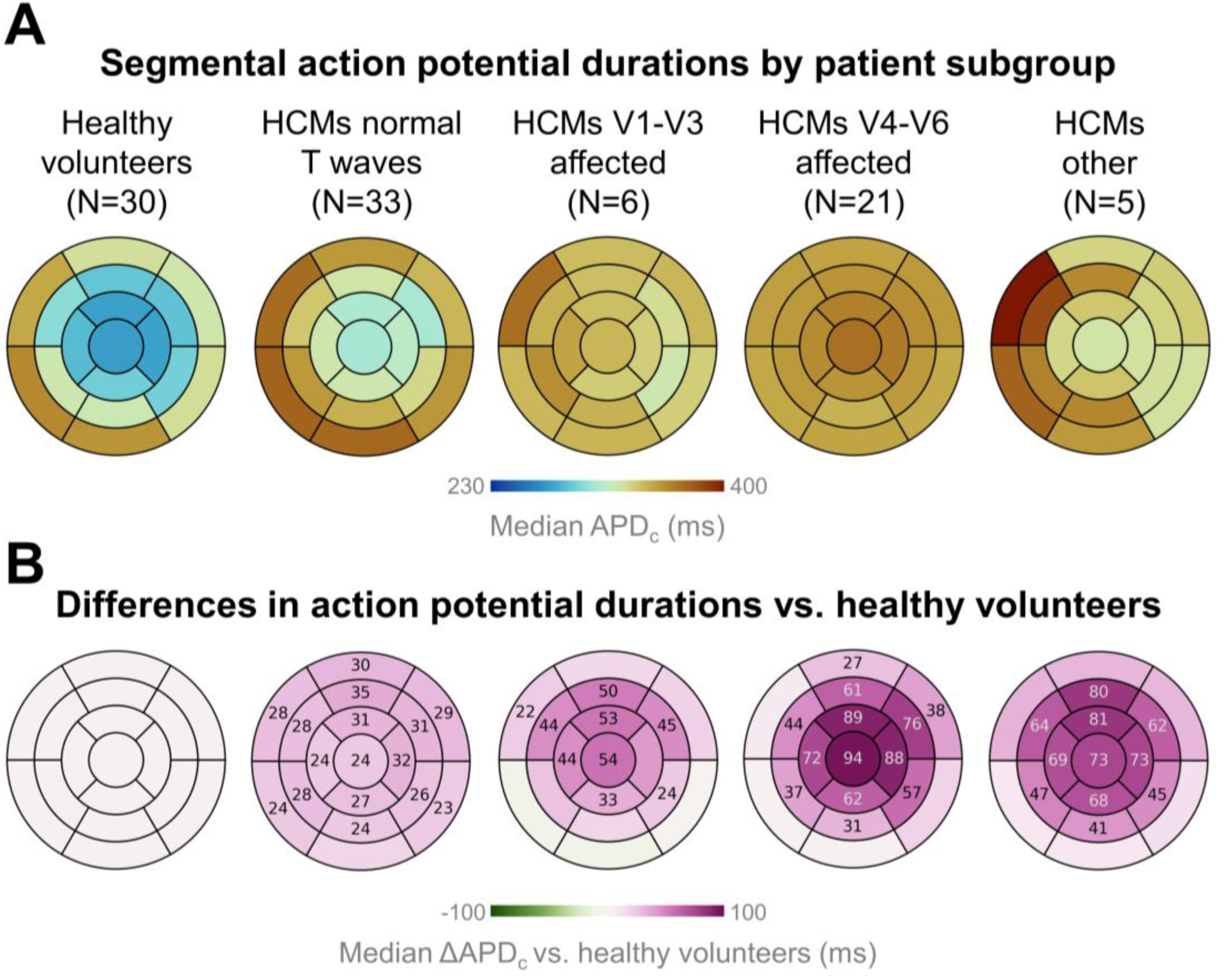
Analysis of HCM repolarisation patterns vs. healthy volunteers. (A) Bullseye plots of median segmental APD_c_ across patients, and (B) bullseye plots of median segmental difference in APD_c_ vs. healthy volunteers (ΔAPD_c_), with annotations in statistically significant segments. Results shown for (from left-to-right): healthy volunteers, HCM patients with normal T-waves, HCM patients with abnormal T-waves in V1-V3, HCM patients with abnormal T-waves in V4-V6, and HCM patients with other T-wave abnormalities.

Healthy volunteers (e.g., Fig. 3A) exhibited pronounced apex-to-base APD_c_ gradients of 60 ms (40-80), with longer APD_c_ at the ventricular base, underlying healthy upright T-waves on the 12-lead ECG. The magnitude of these apex-to-base repolarisation gradients is consistent with those reported previously in ECG imaging studies (Ramanathan et al., 2006). A similar apex-to-base APD_c_ gradient of 60 ms (30-80) was also observed in HCM patients with normal upright T-waves (e.g., Fig. 3B), suggesting that physiological repolarisation patterns may be preserved in some patients despite the presence of structural disease.

Conversely, HCM patients with T-wave abnormalities in leads V1-V3 (e.g., Fig. 3C) showed disruption of the apex-to-base gradient due to patches of prolonged APD_c_ in the apical-to-mid anterior region, in broad agreement with the distribution of affected anterior ECG leads. Finally, HCM patients with V4-V6 affected (e.g., Fig. 3D), the most prevalent abnormal T-wave phenotype, exhibited near-flat or reversed apex-to-base APD_c_ gradients (shorter APD_c_ at the base, with marked prolongation towards the apex).

Statistical comparisons of segmental APD_c_ values between HCM subgroups and healthy volunteers (Fig. 4B) demonstrated that distinct patterns of regional APD_c_ prolongation underlie these abnormal ventricular repolarisation distributions and their associated ECG phenotypes. Although both healthy volunteers and HCM patients with normal T-waves exhibited an apex-to-base APD_c_ gradient, the latter group showed modest but widespread APD_c_ prolongation across most ventricular segments. This likely reflects their mildly prolonged global repolarisation, as indicated by a longer QT_c_ interval of 427 ms (406-433) vs. 408 ms (392-424) in the healthy volunteers.

HCM patients with V1-V3 involvement demonstrated APD_c_ prolongation predominantly in mid-anterior segments and the apex, with the largest difference in the apical segment (ΔAPD_c_ vs. healthy: 54 ms; 95% CI: 23-84 ms), resulting in a flattened apex-to-base gradient. In contrast, HCM patients with V4-V6 involvement exhibited marked APD_c_ prolongation throughout all mid-ventricular and apical segments, again maximal at the apex (ΔAPD_c_ vs. healthy: 94 ms; 95% CI: 74-120 ms), alongside contiguous anterolateral prolongation. This suggests that abnormal T-waves in lateral ECG leads are associated with severe mid-ventricular and apical electrophysiological remodelling sufficient to reverse physiological repolarisation gradients. Finally, HCM patients with other T-wave abnormalities also showed APD_c_ prolongation in mid-ventricular and apical regions, albeit to a lesser extent than the V4-V6 subgroup, with the apex remaining significantly affected (ΔAPD_c_ vs. healthy: 73 ms; 95% CI: 32-120 ms). Examples of HCM cases with other T-wave abnormalities are shown in Supplementary Fig. S3.

Notably, these mid-apical patterns of APD_c_ prolongation did not spatially mirror the distribution of hypertrophy, which was predominantly basal anterior and anteroseptal. Nevertheless, HCM patients with abnormal T-waves exhibited greater maximal LV wall thickness than those with normal T-waves (21±6 vs. 17±5 mm; p=0.002). Notably, whilst almost all HCM patients with any apical hypertrophy had abnormal T-waves and mid-apical APD_c_ prolongation, only one third of HCM patients with abnormal T-waves had apical hypertrophy (Supplementary Figure S8). This suggests that structural remodelling at the apex is more closely linked to APD_c_ changes, unlike typical basal anterior and septal structural remodelling seen in HCM.

Repolarisation inference repeated across the full cohort at stronger regularisation showed high consistency with the inferences performed using tuned regularisation, with a Spearman’s correlation agreement for RT_c_ values of *r* = 0.76 (0.68-0.83) (Supplementary Fig. S6). As expected, stronger regularisation led to the exclusion of more subjects due to weaker T-wave match (11 of 101); however, the full statistical analysis under this setting yielded consistent results (Supplementary Fig. S7, as compared with Fig. 4).

Altogether, these analyses provide evidence that regional prolongation of APD is the primary mechanism underlying abnormal ECG repolarisation patterns in HCM. These findings are consistent with in vitro findings in hypertrophied ventricular cardiomyocytes (Coppini et al., 2013), and align with the broader understanding that LV hypertrophy is associated with APD prolongation (Wolk, 2000).

### 3.5 Regionally prolonged APDc can occur in HCM in the presence of a normal QTc

Despite the considerable regional prolongation of APD_c_ observed in the 32 HCM patients with abnormal T-waves, only three of these individuals had a QT_c_ interval exceeding 480 ms. This is particularly notable given that patients in the V4-V6 T-wave abnormality subgroup had median apical APD prolongation approaching 100 ms (ΔAPD_c_ vs. healthy: 95 ms; 95% CI: 74-120 ms). Previous computational studies have suggested that QT_c_ should increase in proportion to APD prolongation in HCM (Coleman et al., 2024a). By contrast, the present findings suggest that QT_c_ prolongation is not an inevitable consequence of APD_c_ prolongation when this remodelling is regionally confined, highlighting a key limitation of global indices of repolarisation such as the QT_c_ interval, which may fail to detect spatially heterogeneous changes.

Mechanistically, this can be explained by masking effects arising from the intrinsically longer APD_c_ at the ventricular base (Fig. 5). HCM patients with preserved apex-to-base APD_c_ gradients and upright T-waves typically showed basal APD_c_ values around 60 ms longer than those at the apex (Fig. 5A). In such cases, the QT_c_ interval predominantly reflected overall ventricular repolarisation time, which was determined by relatively prolonged basal repolarisation (APD_c_=370 ms in Fig. 5A), rather than apical repolarisation (APD_c_=310 ms). In HCM patients with apical APD_c_ prolongation, the degree of apical remodelling first had to exceed the pre-existing long basal APD_c_ before influencing the QT_c_ interval. For instance, in Fig. 5B, an HCM patient with T-wave abnormalities in V4-V6 showed marked apical APD_c_ prolongation (APD_c_=380 ms); however, this remained comparable to the basal endocardial APD_c_ (360 ms), and the resulting QT_c_ of 430 ms remained within normal range. By contrast, in Fig. 5C, a prolonged QT_c_ interval in a patient with T-wave inversions is attributable to severe apical APD_c_ prolongation (up to 440 ms), which significantly exceeded basal APD_c_ (360 ms) and manifested as a QT_c_ of 490 ms. Collectively, these findings emphasise the limitations of QT_c_ as a sole marker of repolarisation abnormality and highlight the importance of spatially resolved repolarisation metrics.

**Figure 5.**
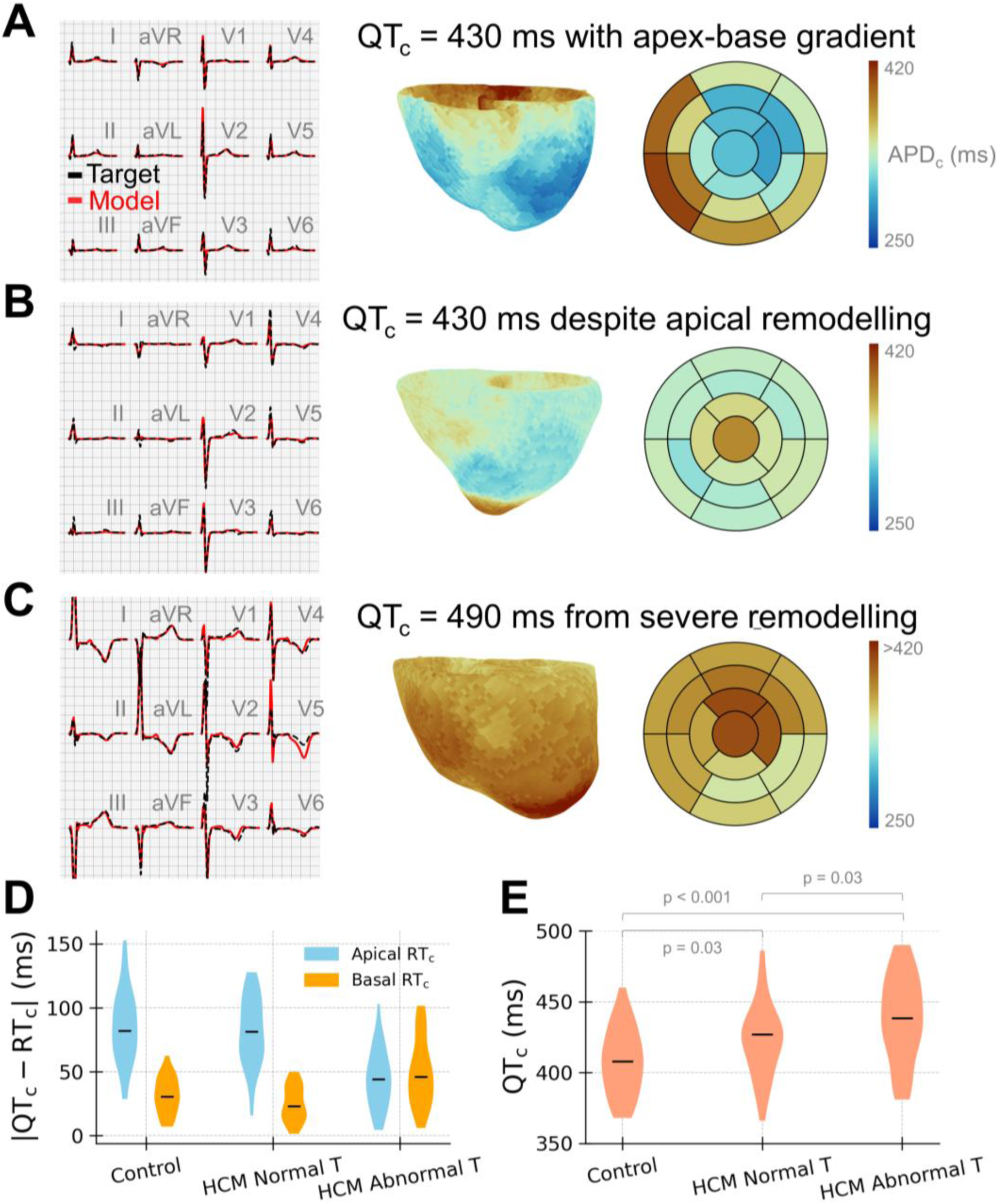
Regional APD prolongation does not necessarily result in proportional QT prolongation. (A–C) Patient 12-lead ECGs and inferred ECGs (left), inferred distributions of ventricular APD_c_ values shown from an anterior view (middle), and corresponding inferred APD_c_ bullseye plots (right). Shown are: (A) HCM patient with normal upright T-waves, normal QT_c_ interval, and a typical apex-to-base APD_c_ gradient; (B) HCM patient with T-wave abnormalities in V4-V6, with marked apical APD_c_ prolongation but yet normal QT_c_ due to masking by prolonged basal APD_c_; and (C) a different HCM patient with T-wave abnormalities in V4-V6 with QT_c_ exceeding 480 ms as a result to mid-apical APD_c_ prolongation exceeding basal APD_c_. (D) Violin plots of absolute difference between QT_c_ and RT_c_ (compared between apical and basal segments), and (E) violin plots of QT_c_, shown for healthy volunteers, HCM patients with normal T-waves, and HCM patients with abnormal T-waves.

The relative contributions of basal and apical repolarisation to the QT_c_ interval were further quantified by computing differences between QT_c_ and RT_c_ (|QT_c_ - RT_c_|) separately for basal and apical segments (Fig. 5D). In healthy volunteers, |QT_c_ - RT_c_| was lower in basal segments (30 ms; 20-40) than in apical segments (80 ms; 70-100), indicating that prolonged basal repolarisation best explained QT_c_ intervals. A similar pattern was observed in HCM patients with normal T-waves, who exhibited similar apex-to-base repolarisation gradients, with |QT_c_ - RT_c_| values in basal segments of 20 ms (20-40) vs. 80 ms (60-100) in apical segments. Conversely, HCM patients with abnormal T-waves showed comparable |QT_c_ - RT_c_| in basal (50 ms; 30-60) and apical (40 ms; 30-60) segments, consistent with mixed apex-base contributions to the QT_c_ interval due to the presence of prolonged apical repolarisation in this subgroup. Interestingly, these HCM patients with abnormal T-waves also manifested slightly longer QT_c_ intervals (439 ms; 422-459) compared with healthy volunteers (408 ms; 392-424) and HCM patients with normal T-waves (427 ms; 406-433) (Fig. 5E). This suggests that mid-apical electrophysiological remodelling can result in a modest increase in QT_c_, even if the full extent of APD_c_ prolongation is not reflected by this global measure.

## 4. Discussion

This study aimed to investigate the mechanistic basis of different repolarisation abnormalities on the 12-lead ECG in HCM, with particular emphasis on understanding how T-wave inversion can occur in the absence of QT_c_ prolongation. This is the first study to use personalised simulation-based inference at scale to reconstruct patient-specific ventricular repolarisation patterns in HCM from standard ECG and CMR clinical inputs. The principal findings are that: (i) distinct ventricular patterns of APD prolongation explain different T-wave abnormalities in HCM; and (ii) regional APD prolongation can occur without significant QT_c_ prolongation, owing to masking by intrinsically longer APDs at the ventricular base.

Consistent with the present findings, APD prolongation has previously been demonstrated *in vitro* in human adult cardiomyocytes from HCM patients (Coppini et al., 2013). However, such studies were limited to septal tissue obtained from surgical myectomies and therefore could not characterise the spatial distribution of APD prolongation across the ventricles in HCM. In contrast, the present study identifies APD prolongation not only in mid-to-apical septal regions, consistent with surgical evidence, but also across other mid-to-apical ventricular segments. This suggests that ionic remodelling in HCM may be spatially widespread rather than confined to the basal septum. Coppini et al. also reported cellular APDs to be 140 ms longer than their controls at 1 Hz pacing, a larger effect than observed here. This discrepancy may reflect the increased severity of disease in their cohort, where almost 50% of their HCM patients had QT_c_ intervals exceeding 480 ms, with all patients requiring surgical relief of longstanding LV outflow tract obstruction. Additionally, isolated cardiomyocytes may exhibit more pronounced APD abnormalities than ventricular tissue, owing to damage caused by isolation and because electrotonic effects act to attenuate spatial differences in repolarisation.

Support for regionally prolonged repolarisation in HCM also comes from ECG imaging studies, where patches of long activation-recovery intervals have been reported in an apical pattern alongside negative apical T-waves (Chow et al., 2024). This aligns closely with the present findings, where prolonged apical APD was also observed also at the epicardial surface (Fig. 3D). Another recent ECG imaging study reported prolonged activation-recovery intervals in HCM, but without segmental analysis or direct correlation with ECG phenotypes (Joy et al., 2024). Extending these epicardial ECG imaging findings, the present study reconstructs electrophysiological substrates throughout the entire myocardial wall, including the interventricular septum, which accounts for majority of the pathological phenotype in HCM.

Further mechanistic support is provided by computational modelling studies. Simulations involving apical-predominant ionic remodelling in HCM have been shown to cause lateral lead T-wave inversions, providing a mechanistic rationale consistent with the present findings (Lyon et al., 2018b). Moreover, the apical patterns of APD prolongation identified here also align with mechanisms proposed to underlie T-wave pseudonormalisation in HCM, as shown in recent modelling work (Coleman et al., 2024b). In this context, regional acute myocardial ischaemia, particularly at the apex, can correct abnormally prolonged APDs and transiently normalise inverted T-waves in HCM, a phenomenon supported by recent clinical observations (López-Ibor et al., 2025; Ma et al., 2024). Although ionic remodelling is likely the electrophysiological basis for the initial APD prolongation, the processes driving the remodelling are unclear (Coppini et al., 2013). Apical phenotypes may be particularly informative in addressing this unknown, where T-wave inversion can precede the development of hypertrophy, alongside other early phenotypic features like apical papillary muscle displacement (Filomena et al., 2023).

Although numerous studies have investigated the prognostic significance of QT_c_ interval prolongation and related ECG markers in HCM (Biagini et al., 2016; Debonnaire et al., 2015; Gray et al., 2013; Güner et al., 2020; Ingegerd et al., 2017; Magrì et al., 2017; Norrish et al., 2022; Zhang et al., 2022), the mechanisms driving QT_c_ prolongation remain incompletely understood. Previous work has suggested contributions from both impaired ventricular activation and repolarisation (Johnson et al., 2011), but spatial considerations have not previously been addressed in detail. The present study demonstrates that APD prolongation does not necessarily translate into QT_c_ prolongation on the ECG, helping to explain why QT_c_ is not heavily weighted in current clinical guidelines (Arbelo et al., 2023; Elliott et al., 2014).

Our findings indicate that the impact of APD prolongation on QT_c_ depends not only on the magnitude of repolarisation delay but also on its spatial distribution. Severe apical APD prolongation may remain undetected by the QT_c_ interval because APD at the ventricular base is intrinsically longer and therefore dominates the QT_c_ interval. Conversely, APD prolongation at the ventricular base would be expected to more readily prolong the QT_c_ interval. As such, global repolarisation metrics like the QT_c_ interval may fail to reflect the severity of localised electrophysiological remodelling. This framework provides a mechanistic explanation for the well-recognised discordance between the high prevalence of T-wave inversion and the relatively low prevalence of prolonged QT_c_ in HCM.

Taken together, these findings demonstrate that simulation-based inference offers a powerful complementary framework for non-invasive assessment of patient-specific electrical function using only routinely acquired 12-lead ECG and CMR data. In future, this approach may support improved patient stratification and pharmacologic evaluation, as spatial repolarisation patterns are increasingly recognised as important determinants of arrhythmic risk and therapeutic response (Chauhan et al., 2006; Chrispin et al., 2026; Coleman et al., 2024a; Yamazaki et al., 2022).

## 5. Limitations

The principal limitation of this work is that it shares inherent constraints common to inverse ECG methods. As the inverse problem is ill-posed, the reconstruction of electrophysiological activity requires regularisation to stabilise the solution, as applied here. While this approach is essential, it may limit the complexity of the reconstructed ventricular behaviour. Although we believe that the present methodology is sufficient to capture the dominant, macroscopic patterns of ventricular repolarisation underlying distinct 12-lead ECG phenotypes in HCM, more subtle repolarisation features may lie beyond the spatial resolution of the method.

Relatedly, a small minority of inferred ventricular substrates exhibited patchy repolarisation patterns that did not conform to their subgroup archetype. While subgroup trends were highly consistent, such cases may pose challenges for patient-specific inferences. Given that potential interacting sources of error exist, future work would benefit from more formal uncertainty-quantification approaches to help identify low-confidence inference results and identifiability limits.

Although the inferred ventricular models were able to reproduce different T-wave phenotypes with good accuracy, the inference method does not yet capture variations in ST-segment elevation or slopes, despite ST-segment abnormalities being common in HCM. Future work may therefore consider inferring ventricular resting membrane potentials, particularly in the context of myocardial ischaemia. However, such extensions were not necessary for the inference of regional APDs pursued in the present study.

The present study did not consider all other disease processes that may contribute to or be associated with repolarisation abnormalities in HCM. Future work may therefore investigate the contribution of additional components of the HCM substrate and their relationship to regional repolarisation abnormalities.

Finally, given the clinical heterogeneity of HCM, subgroup analyses would benefit from application to larger cohorts with greater representation of more severe disease phenotypes. Future work will therefore aim to extend this patient-specific functional analysis to large-scale registry datasets with long-term outcomes, such as the Hypertrophic Cardiomyopathy Registry (HCMR) study (Kramer et al., 2015).

## 6. Conclusion

This study demonstrates that abnormal T-waves in HCM arise from distinct regional patterns of APD prolongation and that such electrophysiological remodelling can occur in the absence of significant QT_c_ prolongation. Protracted mid-to-apical repolarisation appears most pronounced in HCM patients with lateral lead abnormalities, identifying a patient subgroup with particularly impaired electrical function. Altogether, this work provides the first demonstration of the feasibility of reconstructing patient-specific ventricular electrophysiological substrates from standard-of-care 12-lead ECGs and CMR data using simulation-based inference.

## Data Availability

The computational tools used to perform the inferences are available at https://github.com/JamesAlecColeman/sim-based-inf-clin. The clinical ECG and CMR data analysed in this study are subject to data access agreements with the clinical collaborators responsible for the cohort and are therefore not publicly available. Summary statistical data and inferred segmental data supporting the findings of this study are available from the corresponding author upon reasonable request.

## Software availability

The computational tools used to perform the inferences are available at https://github.com/JamesAlecColeman/sim-based-inf-clin.

## CRediT authorship contribution statement

**James A Coleman:** Conceptualisation, Data curation, Formal analysis, Investigation, Methodology, Software, Validation, Visualisation, Writing – original draft, Writing – review & editing. **Julia Camps:** Methodology, Writing – review & editing. **Abdallah Hasaballa:** Methodology, Writing – review & editing. **Rina Ariga:** Data curation, Writing – review & editing. **Betty Raman:** Data curation, Writing – review & editing. **Iacopo Olivotto:** Writing – review & editing. **Hugh Watkins:** Data curation, Writing – review & editing. **Alfonso Bueno-Orovio:** Conceptualisation, Methodology, Funding acquisition, Project administration, Resources, Supervision, Writing – review & editing.

## Acknowledgements

JAC, AIH, IO and ABO are funded by the SMASH-HCM project under Innovate UK grant 10110728 and Horizon Europe Programme grant agreement 101137115. JC acknowledges support from a fellowship from “la Caixa” Foundation (LCF/BQ/PI25/12100029). BR is funded by a Wellcome Career Development Award fellowship (302210/Z/23/Z) and acknowledges support from the NIHR Oxford BRC and BHF Oxford CRE (BHF RE/18/3/34214). IO acknowledges additional support from STRATIFY-HF under Horizon Europe grant agreement 101080905. HW’s laboratory is supported by the BHF’s Big Beat Challenge award to CureHeart (BBC/F/21/220106). The authors further acknowledge the use of the University of Oxford Advanced Research Computing (ARC) facility (https://doi.org/10.5281/zenodo.22558).

For the purpose of open access, the authors have applied a Creative Commons Attribution (CC BY) public copyright licence to any Author Accepted Manuscript version arising from this submission.

---

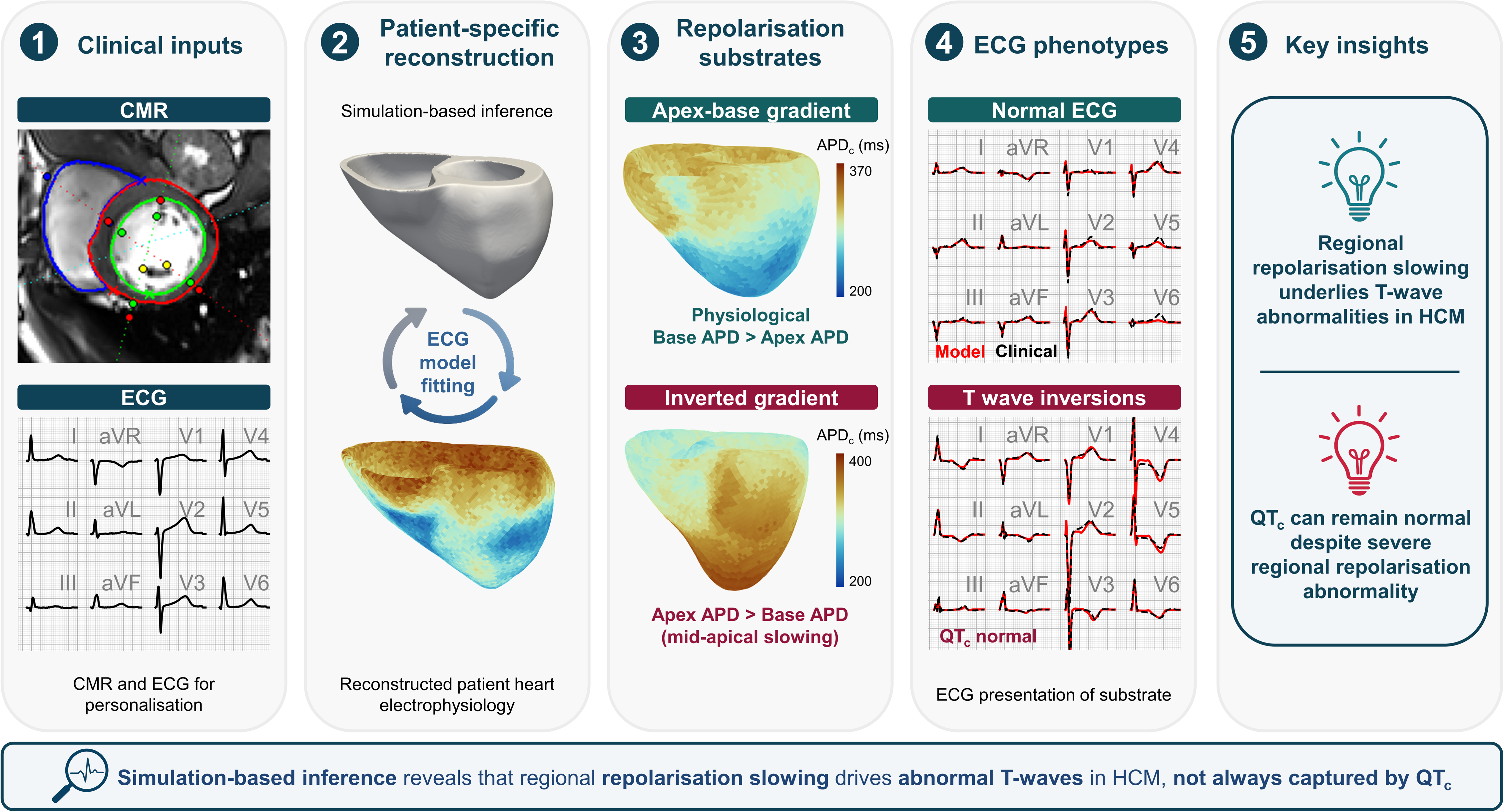

